# S100A9 induces neuroinflammation and aggravates early brain injury after subarachnoid hemorrhage by activating the TLR4/MYD88/NF-κB pathway

**DOI:** 10.1101/2023.03.09.23287076

**Authors:** Guijun Wang, Kesheng Huang, Zhan Zhang, Yujia Guo, Qi Tian, Chengli Liu, Zhijie Li, Zhui Yu, Mingchang Li

**Affiliations:** Department of Neurosurgery, Renmin Hospital of Wuhan University, Wuhan 430060, Hubei Province, China; Department of Critical Care Medicine, Renmin Hospital of Wuhan University, Wuhan 430060, Hubei Province, China; Department of Rehabilitation Medicine, Renmin Hospital of Wuhan University, Wuhan 430060, Hubei Province, China

**Keywords:** S100A9, subarachnoid hemorrhage, neuroinflammation, early brain injury, microglia

## Abstract

**Background:** Subarachnoid hemorrhage (SAH) is a stroke subtype with an extremely high mortality rate, and its severity is closely related to the short-term prognosis of patients with SAH. The S100 calcium-binding protein A9 (S100A9) has been shown to be associated with some neurological diseases, and this study aimed to investigate the relationship between S100A9 and neuroinflammation, as well as its mechanism in SAH.

**Methods:** An enzyme-linked immunosorbent assay (ELISA) was used to detect the concentration of S100A9 in clinical cerebrospinal fluid samples. Furthermore, an in vivo mouse SAH model was established using intravascular perforation; S100A9 knockout mice were used for the in vivo experiments. S100A9 recombinant protein was administered via lateral ventricular injection 1 h before SAH model induction. SAH grade, neurological function score, and brain water content were measured after a specific time. BV2 and HT22 cells and co-culture models were treated with heme chloride to establish an in vitro model of SAH. Paquinimod was used to explore the potential neuroprotective mechanisms of S100A9 inhibition. Western blotting and immunofluorescence staining were used to explore microglial activation, inflammatory responses, and its related protein pathways.

**Results:** The expression of S100A9 protein in the cerebrospinal fluid of patients with SAH increased and was related to the short-term prognosis of patients with SAH; S100A9 was highly expressed in the microglia. S100A9 knockout significantly improved neurological function scores, reduced brain edema, and reduced neuronal apoptosis. S100A9 inhibition with Paquinimod restrained neuronal apoptosis, while administration of recombinant S100A9 aggravated neuroinflammation, activated the TLR4 receptor, promoted NF-κB nuclear transcription, and ultimately aggravated nerve injury.

**Conclusion:** S100A9 protein expression increased after SAH, which induced neuroinflammation and promote neuronal apoptosis by activating the TLR4/MYD88/ NF-κB pathway, ultimately aggravating nerve injury after SAH.

## 1. Introduction

Subarachnoid hemorrhage (SAH) is a type of stroke that accounts for 3–5% of all stroke cases, and is characterized by a high mortality and poor prognosis^1^. Approximately 35% of patients die within 48 h of SAH^2^. The majority of SAH cases are caused by the rupture of an intracranial aneurysm. Based on previous studies, early brain injury (EBI), which refers to the process of pathological changes in brain tissue, is caused within 72 h after SAH, and a significant proportion of survivors develop long-term neurological deficits^3, 4^. There are several key factors leading to poor prognosis after SAH, with EBI being the most studied in recent years^5^. After SAH, the blood-brain barrier (BBB) is destroyed, and neuroinflammation and oxidative stress occur, which are the mechanisms leading to EBI.

Neuroinflammation can occur in the early stages of SAH, and has been shown to lead to neuronal apoptosis^6^. Neuroinflammation with induced neuronal cell death has been classified as one of the main mechanisms underlying secondary brain injury^7^. The pathological processes in brain tissues after SAH are complex, and the underlying mechanisms have been extensively studied, among which the TLR4 pathway is one of the most widely studied pathways^8–10^. The inflammatory response of the body usually takes the form of a pathogen-associated molecular pattern (PAMP) or damage-associated molecular pattern (DAMP), with pattern recognition receptors (PRRs) sensing these patterns and initiating immune responses. One of the most characterized PRR families is the TLR family, and TLR4 has come into focus as a key player in neuroinflammation^11–13^.

Alarmins, also known as DAMPs, are released after macrophage activation and play critical roles in innate immunity. The most prominent members of the DAMP family are heat-shock proteins (HSPs), high mobility group protein B1 (HMGB1), calcium-binding protein S100A9 (also known as myeloid-related protein 14 [MRP-14]), and its binding partner S100A8 (also known as MRP-8)^14^. Neutrophils and monocytes are the most important sources of S100A8 and S100A9, which form stable heterodimers or homodimers, respectively. Both S100A8 and S100A9 have a helix-loop-helix motif with charged amino acid residues, which leads to strong binding to divalent ions. After binding, their conformations change, and these proteins carry-out their respective functions^15^. Extracellular S100A9 interacts with TLR4 and receptor for advanced glycation end products (RAGE), promoting cell activation and recruitment^16^. he S100A9 has been shown to be associated with many neurological diseases; therefore, this study aimed to investigate the relationship between S100A9 and neuroinflammation, and its mechanism after SAH.

## 2. Materials and methods

### 2.1 Human samples

Cerebrospinal spinal fluid (CSF) was collected from patients with SAH; this was approved by the Ethics Review Committee of the Renmin Hospital of Wuhan University (WDRY2021-K070). We collected data from patients hospitalized in our hospital between March 2021 and July 2022, and all patients were between 18 and 75 years of age. Patients with SAH were screened within 24 h of onset of SAH. CSF from a total of 25 patients was collected by lumbar puncture or external ventricular drainage, and consecutive CSF samples were collected from ten of these patients. Normal controls (including, but not limited to, hydrocephalus) underwent lumbar puncture for CSF collection. The CSF was stored on ice and immediately subjected to rotational centrifugation. Supernatants were collected and stored in a refrigerator at −80°C until use for enzyme linked immunosorbent assay (ELISA). All patients with SAH included in the study were scored using the Modified Rankin Scale (mRS) at discharge.

### 2.2 Experimental animals

Wild-type (WT) C57BL/6J male mice (n = 93, 8–10 weeks of age, 22–25 g) and S100A9-KO mice (n = 34, 8–10 weeks of age, 22–25 g) were used in this study. WT mice were purchased from Hunan SJA Laboratory Animal Co.Ltd. S100A9-KO mice were purchased from Cyagen Biosciences (KOAIP210728XW3; Guangzhou, China). The mice used in this study were bred at the Animal Experimental Center of Renmin Hospital, Wuhan University. All mice had ad libitum access to food and water throughout the experiment, and were randomly assigned to groups prior to the start of the experiment. The animal experiments in this study were approved by the Animal Welfare Ethics Review Committee of the Renmin Hospital of Wuhan University.

### 2.3 SAH animal model

An *in vivo* SAH model was established using a modified single-clip method. Anesthesia was induced with isoflurane at a concentration of 5% before the start of surgery, and the mice were intubated with 2–3% isoflurane during surgery (isoflurane was reduced to 1.5% at the time of puncture). The mice were placed in the supine position, and the skin of the neck was dissected after skin preparation, approximately 1.5 to 2 cm in length. The common and internal carotid arteries were then temporarily clipped with an aneurysm clip, followed by the delivery of a thread-containing silicone coating from the external carotid artery to the internal carotid artery, after which the clip was released. The thread was then gradually pushed into the brain along the vessel. When little resistance was detected, it was indicative that the bifurcation of the anterior cerebral artery and the middle cerebral artery has been reached, and pierce was continued for 2 mm. The sham group mice were also subjected to the procedures described above, except for puncturing the vessel upon reaching the bifurcation. Vital signs were monitored every 3 min to ensure that the mice did not experience any distress or death. The entire procedure lasted approximately 10–15 minutes. After recovery from anesthesia, the mice were returned to their cages and were placed in an animal room.

### 2.4 Assessment of SAH grade

All mouse models were graded according to SAH severity. Briefly, the basal cistern was divided into six parts, and each site was scored on a scale of 0 to 3, according to the amount of subarachnoid hemorrhage. All scores were summed to obtain a total score (maximum score of 18). We excluded mice with a total score < 8 from the study.

### 2.5 Experimental design

A total of 127 mice were used in this study, and were subjected to four independent experiments (Fig. 1).

**Figure 1.**
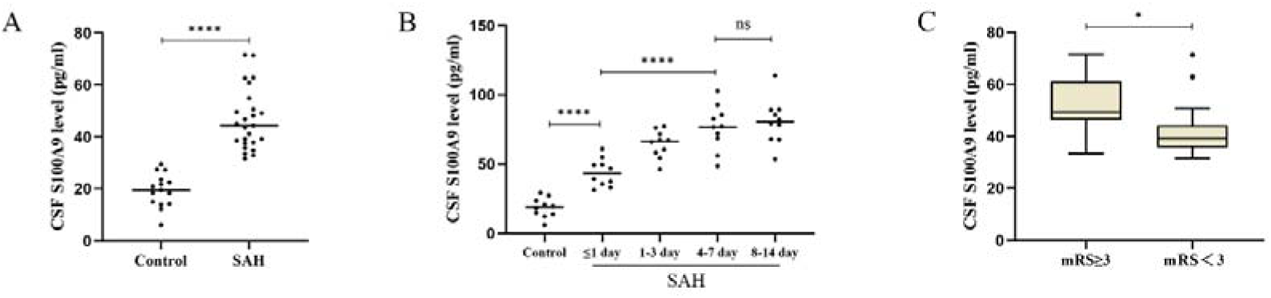
The concentration of S100A9 protein in cerebrospinal fluid (CSF) of patients with SAH, its changes with time and its correlation with short-term prognosis. **A.** The levels of S100A9 were detected by ELISA in CSF samples from control and patients with SAH. **B.** The change trend of S100A9 in CSF of patients with SAH with time. **C.** Horizontal box plot of S100A9 concentration in CSF and mRS at discharge in patients with SAH. *P < 0.05, ****P < 0.0001. Data were represented as the mean ± SD.

#### Experiment 1

To determine the expression of S100A9 protein in the brain tissue of mice and its major effector cells (after SAH), we randomly divided the mice into different time points (3, 6, 12, 24, and 72 h) after SAH induction. Cerebral cortex tissues ipsilateral to the puncture site were collected at corresponding time points for western blotting (n = 3), and the distribution of S100A9 protein was determined by double immunofluorescence staining (n = 3).

#### Experiment 2

To explore the role of S100A9 in the pathological process after SAH, S100A9 knockout mice were used to establish an SAH model. Mice were divided into sham, WT-SAH, and KO-SAH groups. Neurological function of these mice was evaluated at 24 h after SAH (n = 10). The expression of S100A9 protein in knockout mice was verified by western blotting (n = 3). The left and right hemispheres of the mice were collected for assessment of brain water content (n = 3), and hematoxylin and eosin (H&E) staining was performed on the ipsilateral cortex and hippocampus (n = 3).

#### Experiment 3

To explore the effect of S100A9 on neuronal injury after SAH, the mice were divided into three groups: Sham, WT-SAH, and KO-SAH. Neurological function was evaluated 72 h after SAH (n = 6), and neuronal damage was evaluated by TUNEL staining (n = 3) and Nissl staining (n = 3).

#### Experiment 4

To explore whether S100A9 aggravates brain injury after SAH and its effect on microglial expression by inducing neuroinflammation as well as the underlying molecular mechanism, mice were divided into the following five groups: sham, SAH+Vehicle, SAH+S100A9, KO-SAH, and KO-SAH+S100A9. Neurological function was assessed (n=4) 24 h after SAH, and cortical tissues were collected for western blotting (n = 3) and immunofluorescence staining to evaluate the expression of inflammatory factors (n = 3).

### 2.6 ELISA detection

We used ELISA kits to measure S100A9 (HM10001, Bioswamp, China) levels in human CSF samples, including patients with SAH and normal controls. ELISA was performed on all samples in triplicates according to the manufacturer’s instructions.

### 2.7 Intraventricular injection

Intraventricular (i.c.v) stereotactic delivery was performed as described previously^17^. Briefly, intraperitoneal anesthesia was performed with 1% pentobarbital, and the mice were immobilized on a stereoscopic device. A 10 μl microsyringe was inserted into the left ventricle at coordinates of 0.4 mm posterior and 1.0 mm lateral to the prechamber and 3.0 mm inferior to the dural layer. Mouse recombinant S100A9 protein (HY-P74583, MCE, China) was administered at a dose of 100 ng per mouse as previously described^18^. A total volume of 5.0 μL of S100A9 recombinant protein or ddH_2_O was injected 1 h before SAH induction at an injection rate of approximately 0.5 μL/min.

### 2.8 Brain edema measurement

At 24 h after SAH, mouse brains were harvested and weighed to obtain wet weight, dried in a constant-temperature oven for 24 h, and weighed again to obtain the dry weight. The percentage of tissue water content was calculated using the formula ([wet weight − dry weight]/ wet weight) ×100%.

### 2.9 Behavioral analysis

An uninformed experimenter assessed all neurological scores. The modified Garcia score was used to evaluate the degree of neurological damage 24 and 72 h after SAH modeling. Specific scoring criteria were applied as described previously^19^. We also cited another neuroethology to assess the extent of short nerve deficits^20^, but did not create new groupings. In brief, feeding, activity, and deficits were monitored 24 and 72 h after SAH modeling, with a lower score indicative of better neurological function.

### 2.10 Nissl and H&E staining

At 24 or 72 h after SAH, the sacrificed mice were perfused with 4% paraformaldehyde (PFA) and cold phosphate-buffered saline (PBS). The whole brain was then harvested and stored in PFA solution for 24 h. Mouse brains were then cut into 5 µm sections at the indicated sites after embedding in paraffin. Finally, Nissl staining was performed using 5% cresyl blue, and the samples were observed using light microscopy by an uninformed experimenter. For H&E staining, samples were prepared in the same way as for Nissl staining, distinguished by the staining, where paraffin sections were immersed in H&E.

### 2.11 Immunofluorescence and TUNEL Staining

Mice were transcardially perfused with normal saline and 4% PFA 24 h after SAH. Samples for tissue immunofluorescence were prepared using Nissl staining. For cellular immunofluorescence, we seeded cells in 6-well Transwell plates for co-culture and treated them with hemin to mimic an *in vitro* SAH model.

For immunofluorescence staining of tissues and cells, samples were blocked with 5% fetal bovine serum (FBS) at room temperature for 1 h, then diluted primary antibodies of interest were added and incubated overnight (4°C). On the next day, after washing, the corresponding secondary antibodies were added and the cells were incubated for 1 h at room temperature. The slides were cleaned and sealed with DAPI (4-6-diamidino-2-phenylindole) reagent, followed by automatic fluorescence microscopy. The primary antibodies used were anti-S100A9 (1:100, ab92507, Abcam, USA), anti-iba1 (1:200, ab283319, Abcam, USA), anti-GFAP (1:400, ab279289, Abcam, USA), anti-NeuN (1:400, ab104224, Abcam, USA), anti-NF-κB p65 (1:300,80979-1-RR, Proteintech, China), anti-IL-1β (1:200, PAB45925, Bioswamp, China), and anti-TNF-a (1:200, PAB45923, Bioswamp, China). The secondary antibodies were Alexa 488-conjugated and Alexa 594-conjugated (1:400 for S100A9, GFAP, NeuN, TNF-α, IL-1β, and p65; ab150080, ab150077, ab150116, ab150113, Abcam, USA), respectively.

TUNEL staining was performed to determine the degree of apoptosis. TUNEL staining was performed according to the manufacturer’s instructions (G1502, Servicebio, China). After TUNEL labeling, the samples were counterstained with DAPI to detect nuclei; apoptotic nuclei were stained red.

### 2.12 *in vitro* SAH Model

Murine BV2 and HT22 cells were cultured in Dulbecco’s Modified Eagle Medium (DMEM; Procell, China) with 10–15% FBS (Gibco, USA) and 1% penicillin/streptomycin (Gibco, USA).

We stimulated the cells with hemin chloride (Sigma-Aldrich, MO, USA) to mimic the SAH model *in vitro*^19^. Hemin was used to treat BV2 cells, and the subsequent experimental conditions were determined using different treatment times and concentrations. Cells were treated with S100A9 inhibitors (HY-100442, Paquinimod/ABR25757, MCE, China) to mimic an *in vitro* knockout model. Co-cultured cells were randomly assigned into three groups: control, Hemin+Vehicle, and Hemin+Paquinimod. Cells were treated with paquinimod at the same time points as hemin.

### 2.13 Cell Viability Assay

A cell counting kit-8 (CCK-8; Biosharp, China) was used to detect the viability of BV2 cells. One thousand cells were added to each well of a 96-well plate, and the CCK-8 reagent was mixed with the medium at a ratio of 1:9 to form a working solution. 100 μL of working solution was added to each well and incubated for 1 h in a cell incubator. Cell viability was measured at 12 h after heme or paquinimod induction.

### 2.14 Flow Cytometry

Flow cytometry was performed to assess neuronal death using an Annexin V PE/7-AAD kit (#CA1030, SolarBio, China). The data were analyzed using Novocyte Express (Santa Clara, CA, USA). The dead cell count was calculated using the following formula: [number of Annexin V PE+/7-AAD+ cells/number of total cells] × 100%.

### 2.15 Western Blot Analysis

Brain tissue was weighed and lysed for protein extraction. Subsequently, the concentration of each sample was determined using the BCA method, followed by normalization. The extracted proteins were subjected to gel electrophoresis and the bands were transferred to a polyvinylidene difluoride (PVDF) membrane which was then washed and blocked in a 5% bovine serum albumin solution for 1 h. The membrane was then incubated with diluted primary antibody solution at 4°C overnight. The next day, the membrane was washed three times and incubated with the appropriate secondary antibodies for 1 h at room temperature. Membrane bands were detected using an enhanced chemiluminescence (ECL) reagent kit (BL520B, Biosharp, China) after washing three times, and images were acquired using the ChemiDoc™ Touch Imaging System (BIORAD, USA). Images were analyzed using the ImageJ software (Image J 1.4, NIH, USA). The primary antibodies included anti-S100A9 (1:800, ab92507, Abcam, USA), anti-iba1 (1:1000, ab283319, Abcam, USA), anti-IL-1β (1:200, PAB45925, Bioswamp, China), anti-TNF-a (1:200, PAB45923, Bioswamp, China), anti-TLR4 (1:1000, PAB47910, Bioswamp, China), anti-MYD88 (1:1000, PAB47936, Bioswamp, China), anti-NF-κB p65 (1:3000,80979-1-RR, Proteintech, China), anti-phosphorylated NF-KB p65 (1:1000, 3033S, CST, China), and anti-GAPDH (1:1000, GB15002, Servicebio, China). The secondary antibodies included HRP-labeled goat anti-rabbit (1:20,000; B0047, boerfu, China) and HRP-labeled goat anti-mouse (1:20,000; B0048, boerfu, China) antibodies.

### 2.16 Statistical analysis

Statistical analyses were conducted with SPSS 19.0 software (IBM Corp.) and GraphPad Prism 9.0 software (GraphPad Software, Inc.). Data from the experiments were presented as the mean ± SD. For data conforming to normality, we analyzed the significant differences among groups using the Student’s t-test. A nonparametric test or Kruskal–Wallis test was used to analyze the significant differences among groups in which the data failed to achieve normality. P < 0.05 was considered statistically significant.

## 3. Results

### 3.1 The level of S100A9 in CSF is closely related to the prognosis of patients with SAH

To determine whether there was any difference between S100A9 protein levels in the CSF of patients with SAH and normal controls, we selected a total of 25 patients with SAH with onset time within 24 h and 15 normal controls. We collected the first CSF sample at admission, for ELISA, and the results showed that the concentration of S100A9 protein in the CSF of patients with SAH with onset time ≤ 24 hours was significantly higher than that of the control group (Fig 1A, P < 0.0001). To further clarify the changes in S100A9 concentration in the CSF of patients with SAH over time, data from 10 patients were consecutively collected. The results showed that the concentration of S100A9 in the CSF of patients with SAH increased with time after the onset of SAH (Fig 1B), and stabilized after 4–7 days, with no significant difference between days 4–7 and days 8–14 (P > 0.05).

In addition, to understand whether there is a correlation between S100A9 concentration in the CSF and short-term outcomes in patients with SAH, we collected the mRS scores of the previous 25 patients at discharge. We graded the mRS Scores as 0–2 (favorable outcome) and 3–6 (unfavorable outcome). The horizontal box plot showed that the higher the concentration of S100A9 protein in the CSF of patients with onset time ≤ 24 h, the worse the prognosis of the patients at discharge (Fig 1C).

### 3.2 SAH model mortality and grade score

A total of 25 mice were used in the sham-operated group, and 102 mice (68 WT and KO mice) were used to establish SAH models. Finally, 14 mice were excluded (SAH grade score < 8; Fig 2A). The overall mortality rate of the SAH modeling was 24.51% (25/102), and the mortality rates of WT and KO mice were 25% (17/68) and 23.53% (8/34), respectively. A representative schematic of the successful modeling of SAH is shown in Fig 2B. There was no significant difference in the SAH grade scores in all the SAH groups (Fig 2C, P > 0.05).

**Figure 2.**
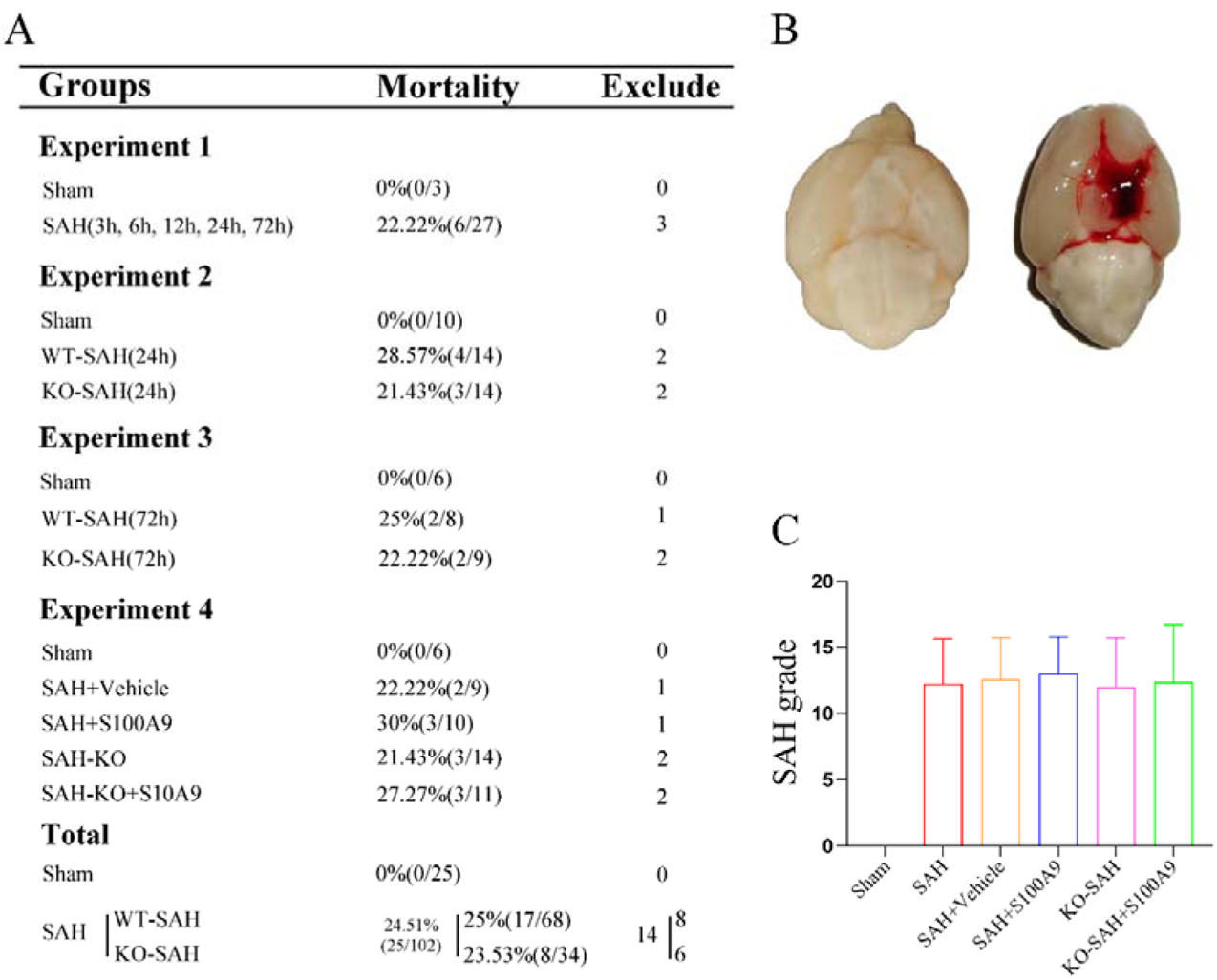
Experimental design, animal groups, mortality, and SAH grading in mice. **A.** Experimental design, animal usage, and mortality. **B.** Representative brain images from the SAH. **C.** SAH grading scores in each group. Data were represented as the mean ± SD.

### 3.3 Temporal Expression of S100A9

The expression of S100A9 protein *in vivo* was detected by western blotting (Fig 3A). The results showed that the expression of S100A9 began to increase at 3 h, reached a peak at 24 h, and then decreased at 72 h (P < 0.05, Fig 3B). Double immunofluorescence staining confirmed that S100A9 mainly colocalized with microglia, but not with astrocytes or neuronal cells, in the cerebral cortex (Fig 3C).

**Figure 3.**
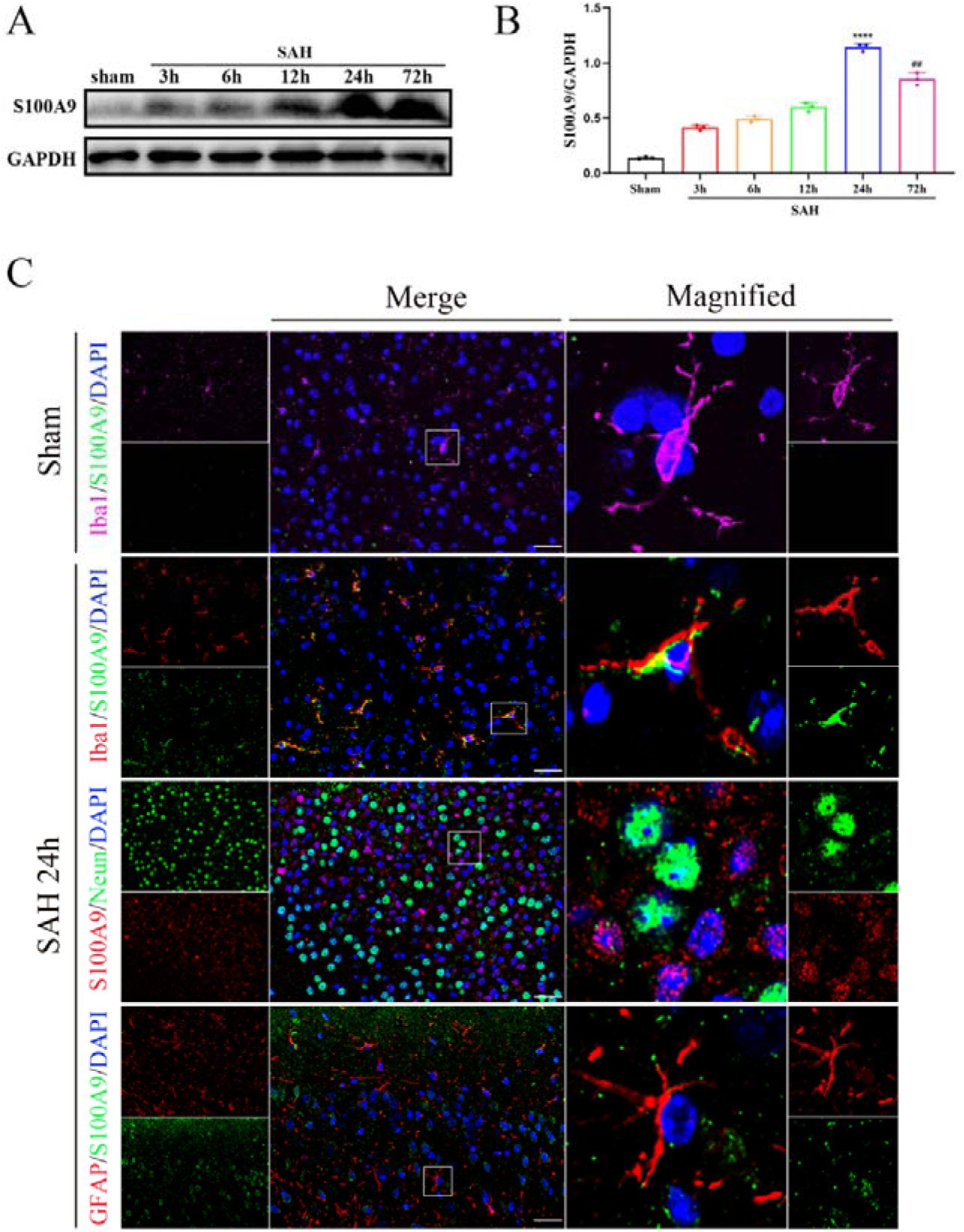
Expression and distribution of S100A9 after SAH in mice. **A-B.** Representative western blotting images and quantitative analyses of S100A9 expression after SAH. n = 3 per group. ****P < 0.0001 vs Sham group; ^##^P < 0.01 vs SAH 24 h group. **C.** Representative images of double immunofluorescence staining for S100A9 in microglia, astrocytes, and neurons at 24 h after SAH. n = 3 per group. Scale bar = 50 mm. Data were represented as the mean ± SD.

### 3.4 S100A9 knockout can improve neurological score after SAH and alleviate EBI

Neurological deficits were assessed using the modified Garcia score and the neurological score 24 h after SAH was analyzed. The neurobehavioral scores changed significantly after SAH compared with the sham group (P < 0.0001, Fig 4A, B), and these changes were reversed by S100A9 knockdown. Brain edema after SAH is closely associated with poor prognosis. The water content of the left and right hemispheres increased significantly 24 h after SAH. However, compared to WT mice, the brain water content of KO mice decreased after SAH (Fig 4C). We confirmed the absence of S100A9 expression in S100A9 knockout mice using western blotting (Fig 4D-E). H&E staining was used for pathological analysis of cortical and hippocampal neurons. The results showed that the number of neurons of the opposite sex increased in the cortex and hippocampus of the mice after SAH, and obvious edema appeared, which was ameliorated by S100A9 knockout (Fig 4F).

**Figure 4.**
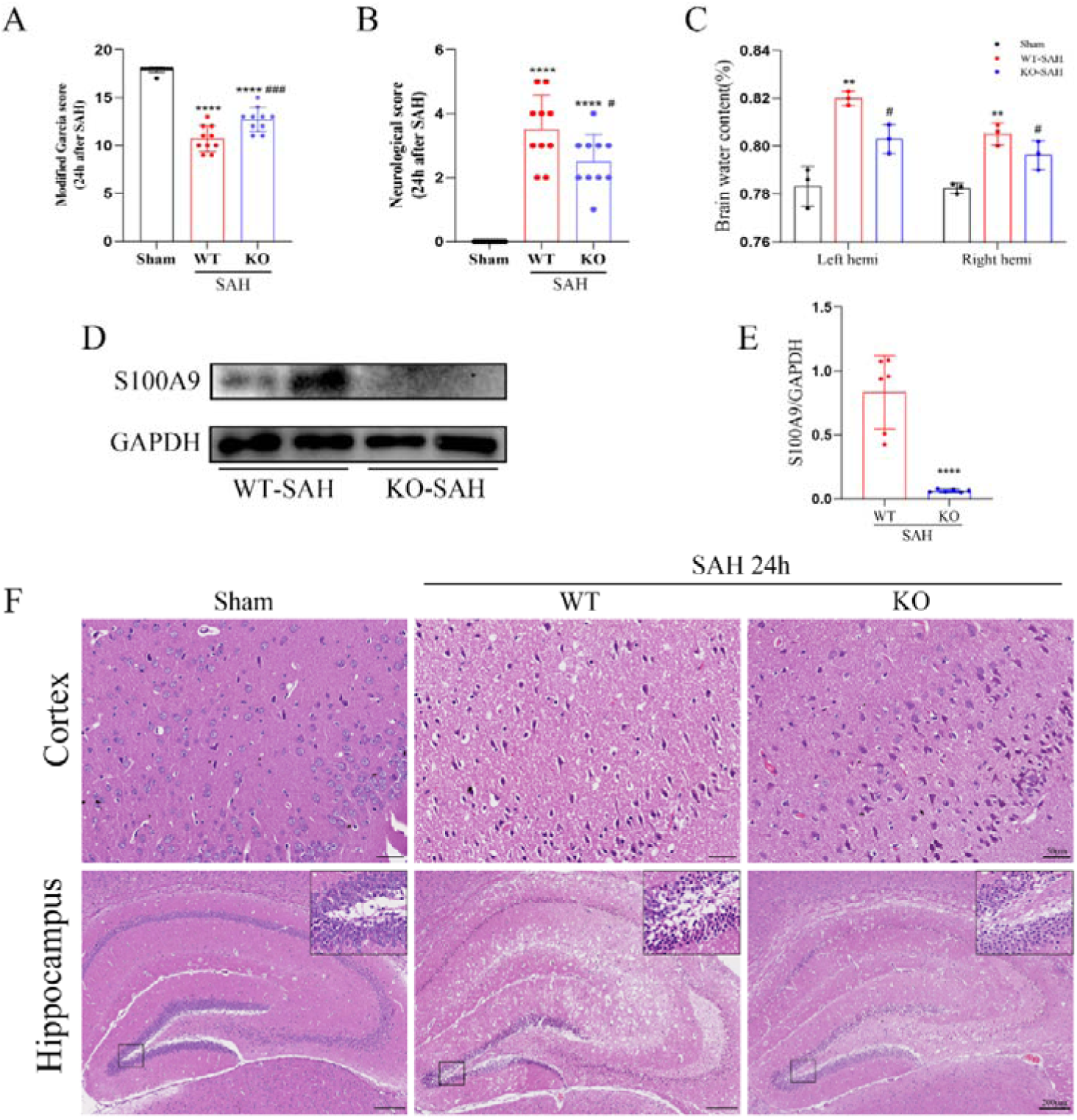
S100A9 knockout can improve neurological deficit, brain edema, and reduce the damage of cortex and hippocampus after SAH 24 h. **A, B.** The modified Garcia and neurological score of each group. n = 10 per group. ****P < 0.0001 vs Sham group; ^###^P < 0.001, ^#^P < 0.05 vs WT-SAH group. **C.** Quantification of brain water content. n = 3 per group. **P < 0.01 vs Sham group; ^#^P < 0.05 vs WT-SAH group. **D, E.** Representative picture and quantitative analysis of western blotting from S100A9 knockout mice. n = 3 per group. ****P < 0.0001 vs WT group. **F.** Representative H&E staining images of brain slides. n = 3 per group. Scale bar = 50/200 µm. Data were represented as the mean ± SD.

### 3.5. S100A9 knockout can improve neuronal apoptosis after SAH

We again used the modified Garcia and neurological scores to evaluate neurological function 72 h after SAH. The results were similar to the changes observed at 24 h (P < 0.0001, Fig 4A, B). TUNEL and Nissl staining were used to observe the degree of neuronal damage in the ipsilateral cortex and hippocampus after SAH. TUNEL staining showed that, compared with the sham group, the number of apoptotic neurons in the cortex of the mice increased after SAH, while S100A9 knockout reduced neuronal apoptosis. The results of Nissl staining showed that the number of neurons decreased mainly in the cortex, corneum area (CA) 1, CA3, and dentate gyrus (DG), and the neurons in these areas showed obvious atrophy 72 h after SAH (Fig 5E–G). These changes were significantly ameliorated by S100A9 knockdown.

**Figure 5.**
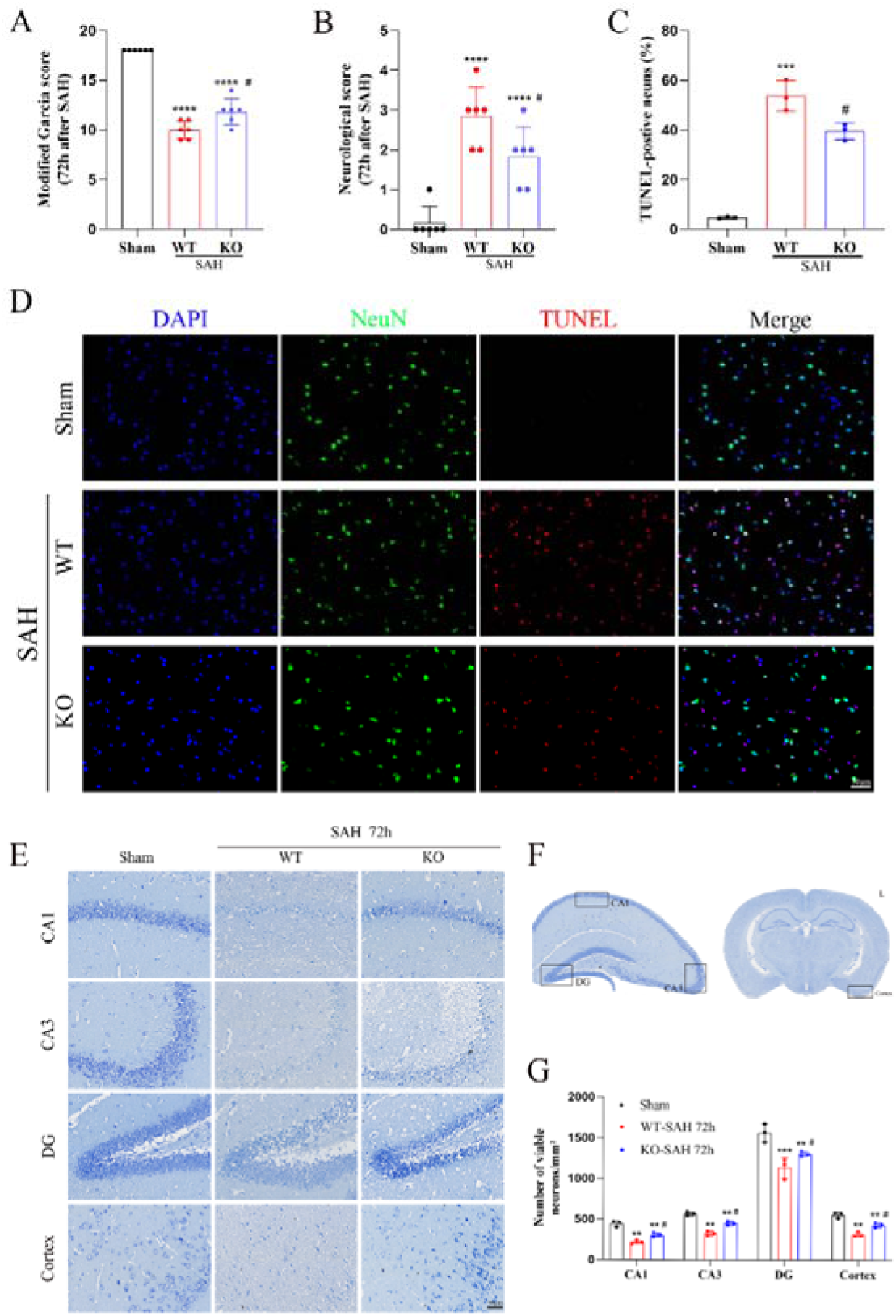
Effect of S100A9 knockout on cortical and hippocampus neuronal injury at 72 h after SAH. **A, B.** The modified Garcia and neurological score of each group. n = 6 per group. ****P < 0.0001 vs Sham group; ^#^P < 0.05 vs WT-SAH group. **C, D.** Representative microphotograph showed the co-localization of TUNEL positive cell (red) with NeuN (green) and quantitative analysis. n = 3 per group. ***P < 0.001 vs Sham group; ^#^P<0.05 vs WT-SAH group. **E–G.** Representative picture of Nissl staining and quantitative analysis. n = 3 per group. ***P < 0.001, **P < 0.01 vs Sham group; ^#^P < 0.05 vs WT-SAH group. Scale bar = 50 µm. Data were represented as the mean ± SD.

### 3.6 Paquinimod reduce neuronal apoptosis in an *in vitro* SAH model

We treated BV2 cells with hemin and detected S100A9 protein expression by western blotting at different time points. At the same time, the effects of hemin and Paquinimod at different concentrations in BV2 cells were detected by CCK8 assay. The results showed that S100A9 expression was highest in BV2 cells treated with hemin for 12 h (Fig 6A). The CCK8 assay results showed that cell viability decreased significantly after adding hemin, and the optimal concentration was 160 μmol/L hemin (cell viability 50.42 ± 4.44%; Fig 6B). In addition, the results also showed that 20 μmol/L Paquinimod (resulting in cell viability 73.37 ± 3.54%) was the optimal treatment concentration after hemin induction of BV2 cells (Fig 6C).

**Figure 6.**
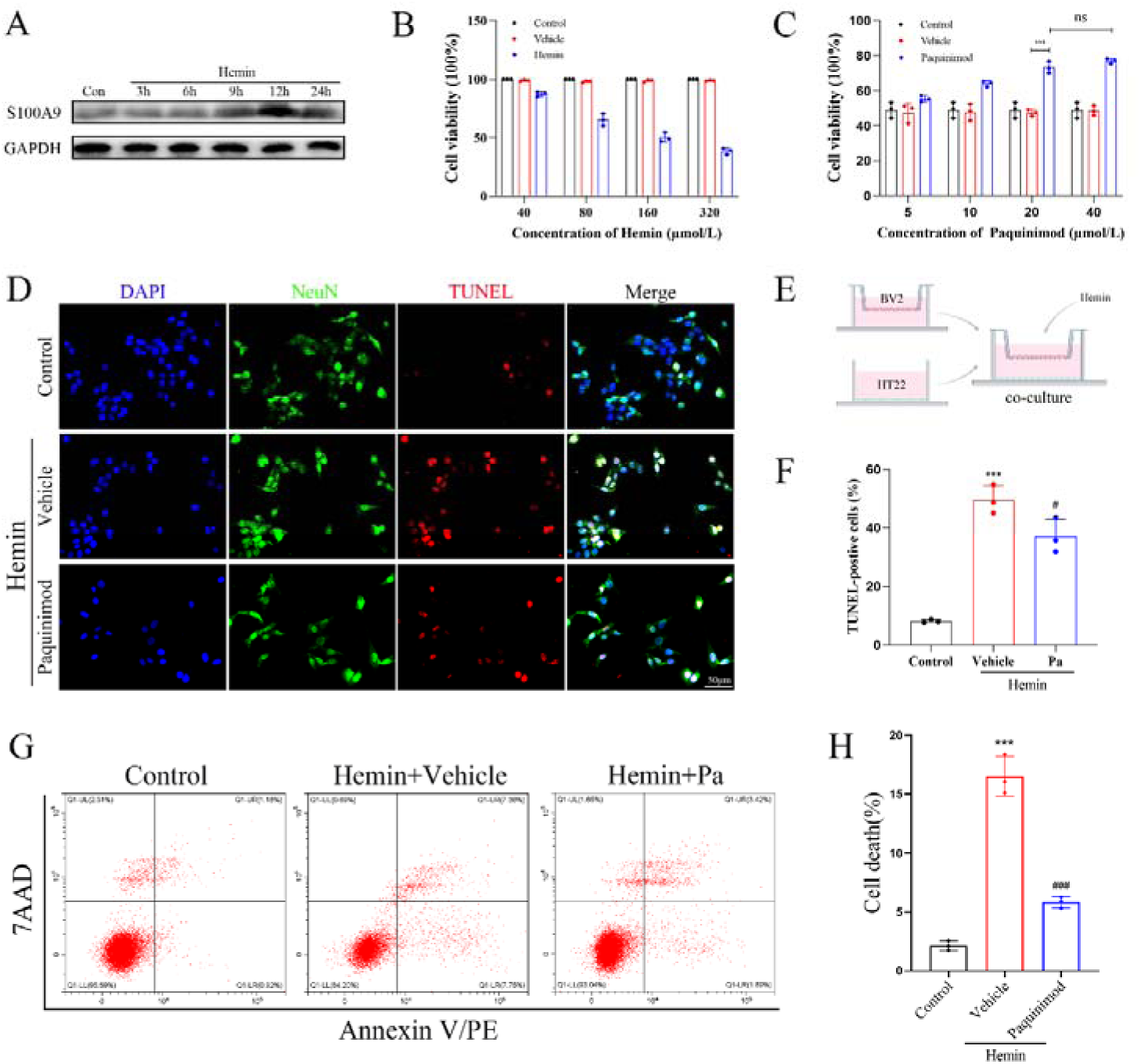
Paquinimod can inhibit hemin-induced neuronal apoptosis. **A.** Representative western blotting picture of S100A9 in hemin-stimulated BV2 cells over time. **B, C.** CCK-8 results showed that the cell viability decreased significantly after treatment with hemin (160 mmol/L). Paquinimod (20 μmol/L) rescued the process. n = 3 per group. ***P < 0.001. **D–F.** Representative microphotograph showed the co-localization of TUNEL positive cell (red) with NeuN (green) and quantitative analysis. n = 3 per group. ***P < 0.001 vs control group; ^#^P < 0.05 vs hemin+vehicle group. **G, H.** Representative flow cytometry images of HT22 cells, apoptotic cells represented by 7-AAD+/Annexin V+ ratio. n = 3 per group. ***P < 0.001 vs control group; ^###^P < 0.001 vs hemin+vehicle group. Scale bar = 50 µm. Data were represented as the mean ± SD.

BV2 and HT22 cells were co-cultured in a hemin-containing medium to explore the effect of S100A9 on the apoptosis of HT22 cells *in vitro* (Fig 6E). Neuronal apoptosis was significantly increased after heme addition compared with that in the control. Paquinimod administration reversed hemin-induced apoptosis (P < 0.05; Fig 6D, F). Flow cytometry confirmed that Paquinimod reversed hemin-induced apoptosis (Fig 6G, H).

### 3.7 S100A9 aggravates nerve injury by inducing neuroinflammation

To further explore the relationship between S100A9 and neuroinflammation, we introduced the mouse S100A9 recombinant protein, and the mice were divided into five groups: Sham, SAH+vehicle, SAH+S100A9, KO-SAH, and KO-SAH+S100A9. Western blotting and immunofluorescence staining were performed to detect the expression of inflammatory factors (TNF-α and IL-1β). The neurobehavioral score results showed that S100A9 administration aggravated neurological damage after SAH in WT mice, and the results were the same in S100A9 knockout mice (Fig 7A, B). The results of western blotting showed that S100A9 recombinant protein administration increased the expression of TNF-α and IL-1β after SAH in the mice, while S100A9 knockout significantly reduced the expression and increased again after S100A9 recombinant protein administration (Fig 7C–F). This result was confirmed by immunofluorescence staining (Fig 7G and H). In addition, we also confirmed that the administration of S100A9 recombinant protein increased the expression of iba1 (Fig 7C), which may also be one of the underlying mechanisms for the increased expression of inflammatory factors.

**Figure 7.**
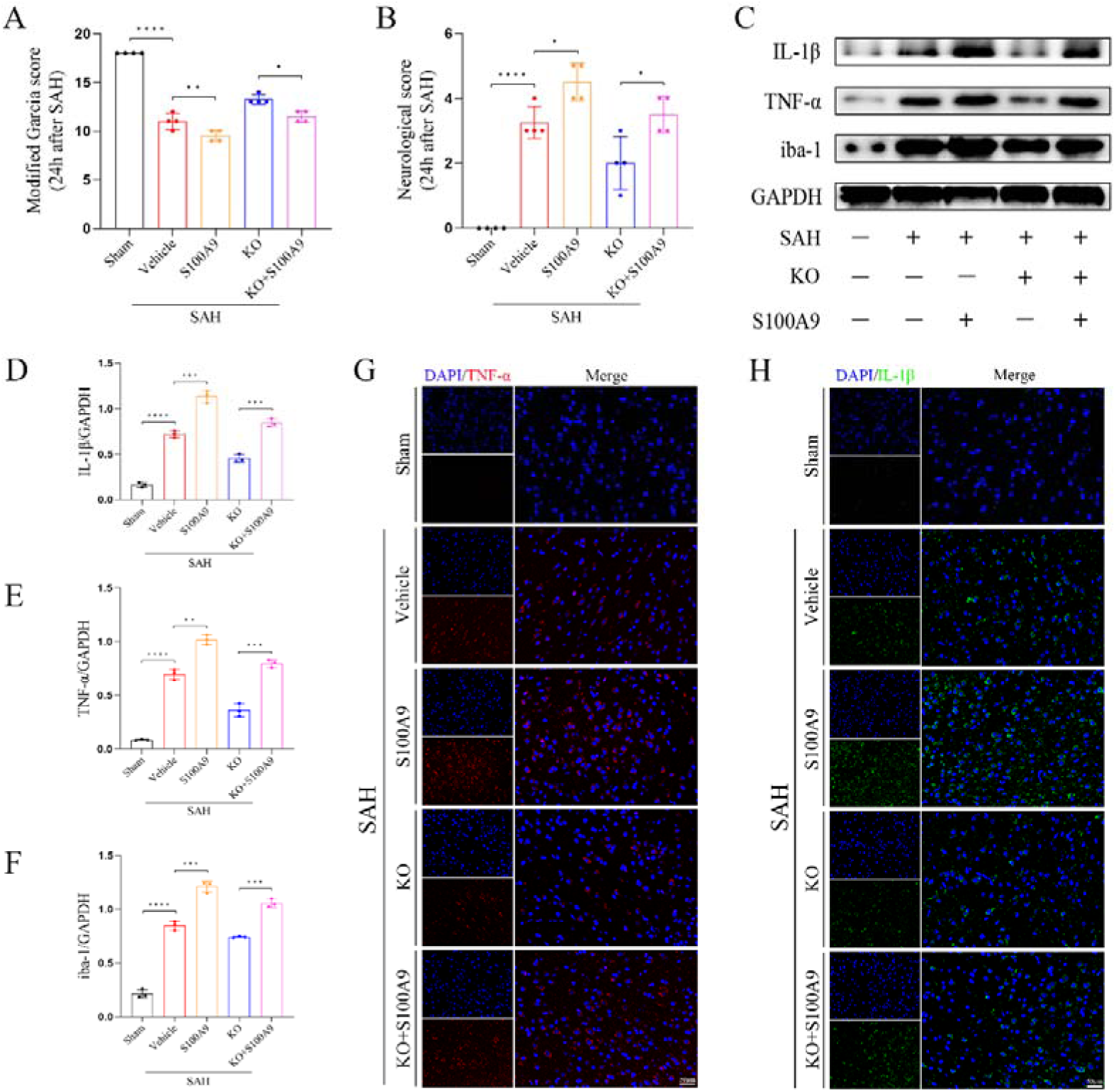
S100A9 aggravates neuroinflammation after SAH. **A, B.** Modified Garcia and neurological scores of each group. n = 4 per group. ****P < 0.0001. **P < 0.01. *P < 0.05. **C–F.** Representative images and quantitative analysis of western blotting for inflammatory factors and microglial markers. n = 3 per group. ****P < 0.0001. ***P < 0.001. **P < 0.01. **G, H.** Representative immunofluorescence images of inflammatory factors. n = 3 per group. Scale bar = 50 µm. Data were represented as the mean ± SD.

### 3.8 S100A9 knockout can attenuate neuroinflammation by inhibiting the TLR4/MYD88/NF-κB pathway

To further explore the potential mechanism by which S100A9 aggravates neuroinflammation, we examined the expression of related pathways in mice after SAH using western blotting. The results showed that the expression of iNOS, TLR4, MYD88, p-IκBα, and p-p65 increased after SAH in mice, and the expression of these proteins was significantly increased after the administration of S100A9 recombinant protein. The expression of these proteins decreased after S100A9 knockout and increased after the reapplication of S100A9 recombinant protein. This suggests that S100A9 knockout inhibits the activation of the TLR4/MYD88/NF-κB pathway and reduces the activation of microglia to alleviate inflammation (Fig. 8A-H).

**Figure 8.**
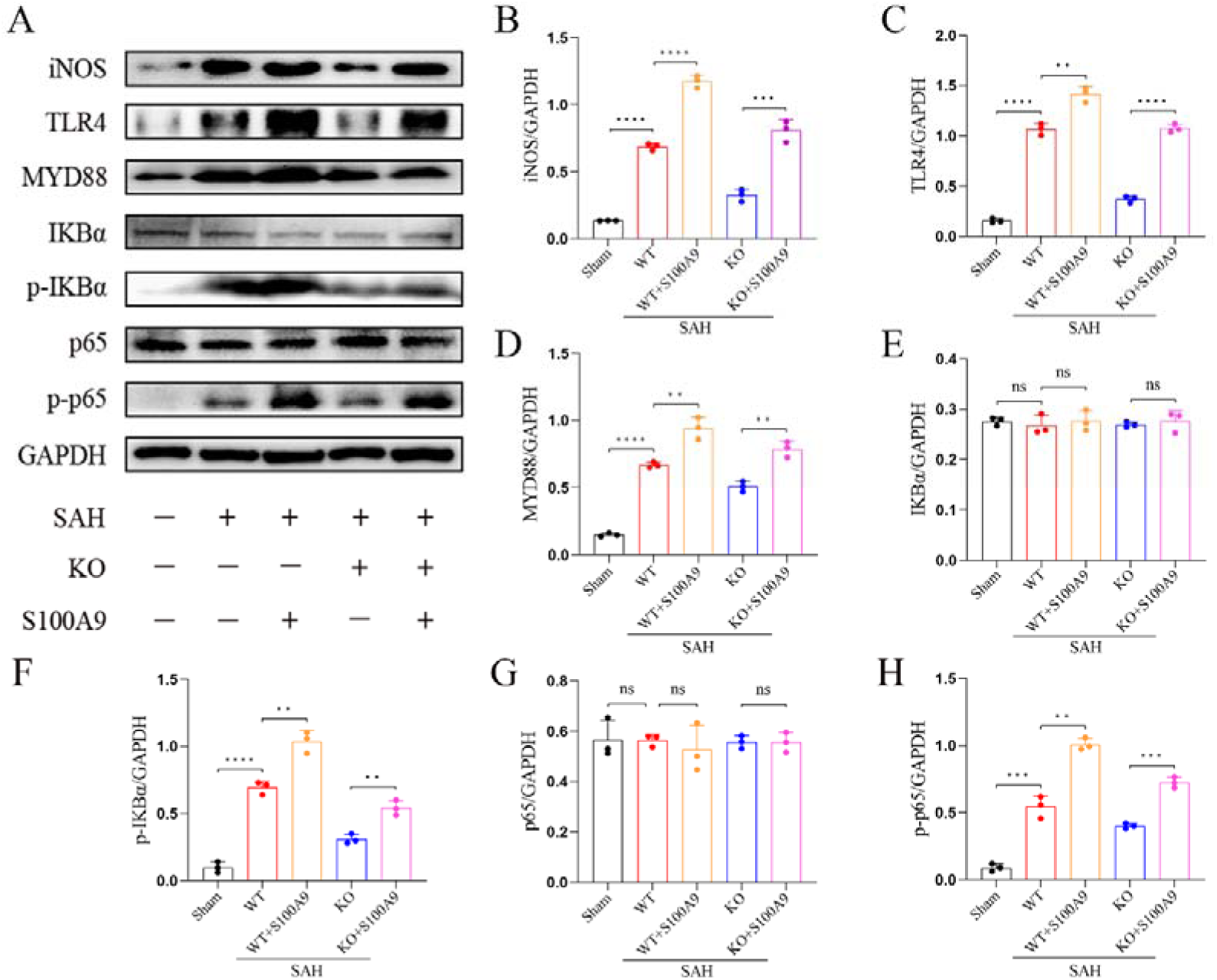
S100A9 induces neuroinflammation by activating the TLR4/MYD88/NF-κB pathway. **A–H.** Western blotting and quantitative analyses of iNOS, TLR4, MYD88, IκBα, p-IκBα, p65, and p-p65 expression. n = 3 per group. ****P < 0.0001. ***P < 0.001. **P < 0.01. Data were represented as the mean ± SD.

### 3.9 Paquinimod reduced the nuclear transcription of NF-κB by inhibiting the TLR4/MYD88 pathway

Similarly, we verified the potential mechanism by which S100A9 aggravates neuroinflammation in an *in vitro* SAH model. Western blotting results showed that the expression of iNOS, TLR4, MYD88, p-IκBα, and p-p65 in BV2 cells increased after hemin stimulation, and their expression continued to increase after the administration of S100A9 recombinant protein. After Paquinimod treatment, the expression of these proteins was significantly decreased compared with that in the untreated group, and the expression was increased after re-administration of S100A9 recombinant protein. Immunofluorescence analysis also verified that Paquinimod treatment inhibited the nuclear transcription of NF-κB after hemin stimulation, while the addition of S100A9 recombinant protein reactivated the nuclear transcription of NF-κB (Fig. 9A-I).

**Figure 9.**
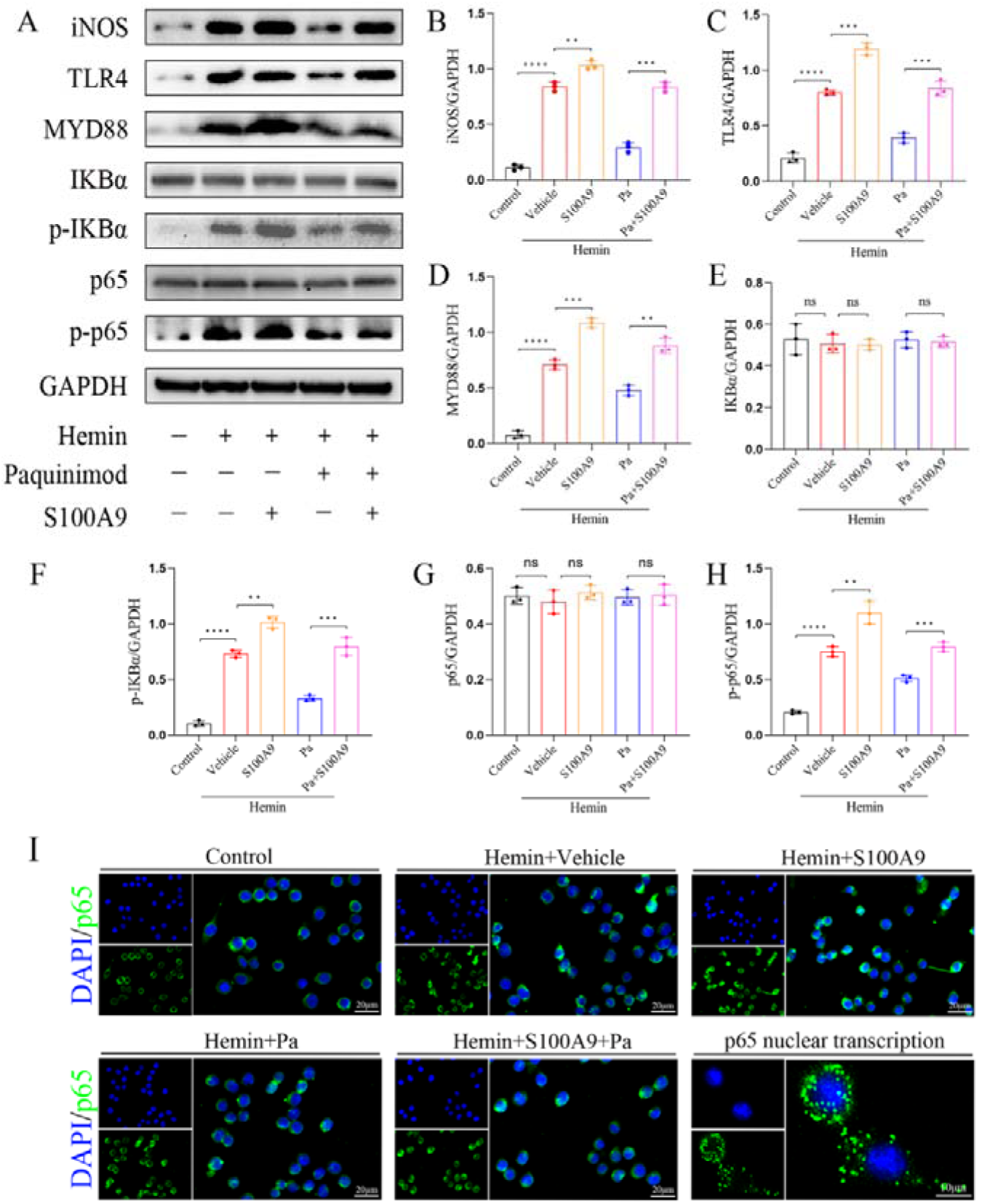
Paquinimod can inhibit the nuclear transcription of NF-κB in *in vitro* SAH model. **A–H.** Western blotting and quantitative analyses of the expression of iNOS, TLR4, MYD88, IκBα, p-IκBα, p65, and p-p65. n = 3 per group. ****P < 0.0001. ***P < 0.001. **P < 0.01. **I.** Representative immunofluorescence images showed that Paquinimod inhibited nuclear transcription in different groups, and representative images of nuclear transcription. n = 3 per group. Scale bar = 50/20 µm. Data were represented as the mean ± SD.

## 4. Discussion

The S100 family proteins were first isolated from the bovine brain as neuroproteins in 1965. S100A8, S100A9, and S100A12 are found at high concentrations in inflamed tissues, with neutrophils and monocytes being the most abundant cell types harboring these proteins^21^. In previous studies, S100A9 was confirmed to be associated with various diseases, in which the central hub is the inflammatory response, including infection-induced inflammation^22^, metabolic inflammation^23^, inflammation caused by immune system dysfunction^24–26^, inflammation caused by degenerative diseases^27–29^, and inflammation-related to neurodegenerative diseases^30^. S100A9 is highly expressed in glial cells, but little is known about its role in many nervous system diseases.

In this study, we established a mouse SAH model and applied S100A9 knockout to explore the role of S100A9 in SAH. In our study, we found that S100A9 knockout mice performed more normally after SAH, and the relevant scores were significantly improved compared to WT mice. This was confirmed by assessment of brain edema and H&E staining after SAH. Our Nissl and TUNEL staining results showed that S100A9 knockout reduced neuronal apoptosis and improved short-term outcome (72 h) in the SAH model. We further verified that S100A9 also aggravated brain injury by inducing an inflammatory response, and further explored that S100A9 mainly activated the TLR4/MYD88 pathway to phosphorylate p65 and nuclear transcription, leading to the activation of microglia and promoting the release of inflammatory factors (Fig 10).

**Figure 10.**
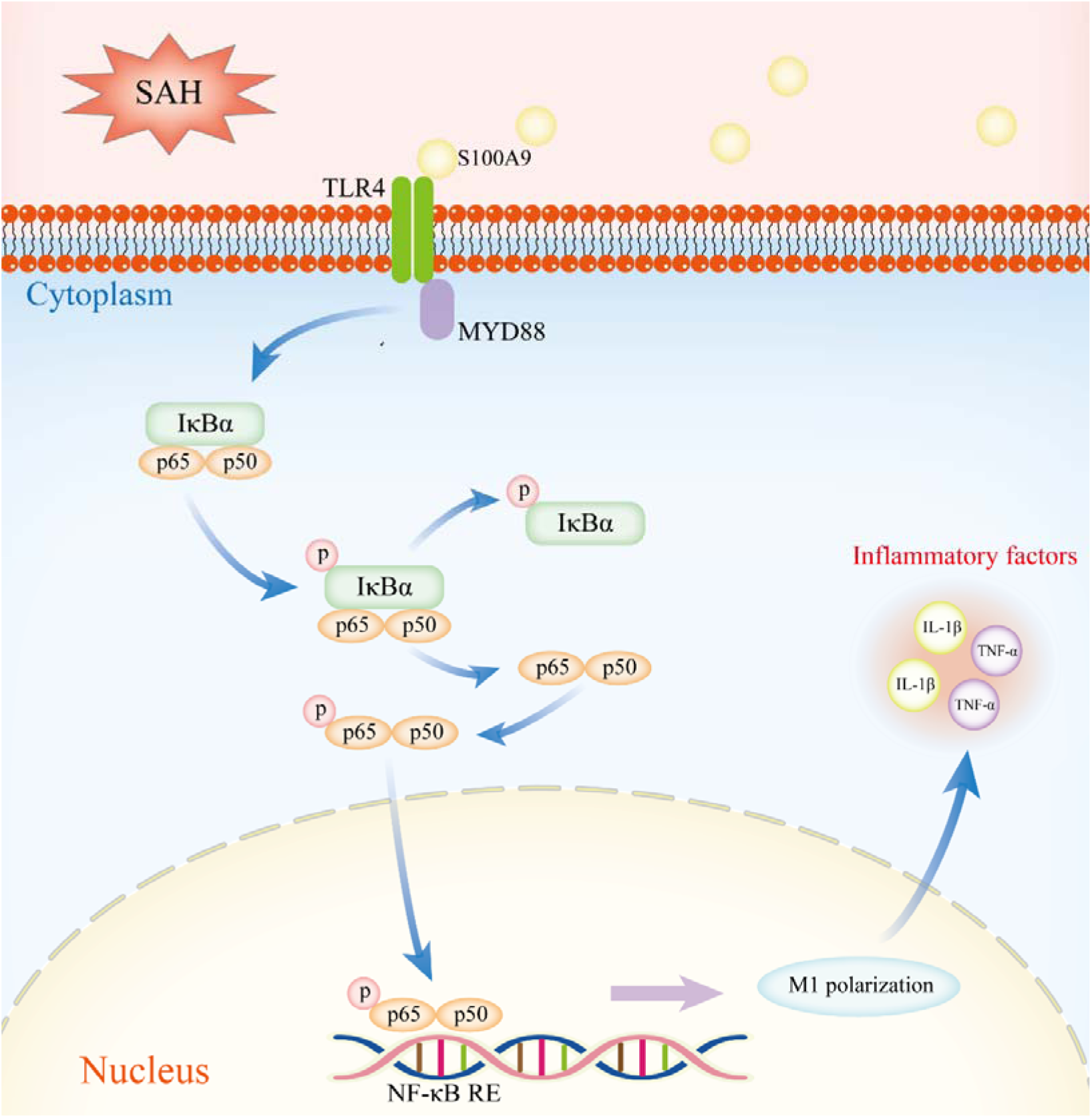
Schematic representation of the mechanism by which S100A9 induces neuroinflammation after SAH via activation of the TLR4/MYD88/NF-κB pathway. The increased expression of S100A9 protein after SAH, activates the TLR4 receptor on microglia, and activates IκBα through MYD88 to release p65. p65 is further phosphorylated into the nucleus and binds to the effector site, inducing the M1 polarization of microglia, releasing inflammatory factors, and ultimately aggravating the early brain injury after SAH.

EBI generally refers to pathological events occurring within 72 h after SAH onset. There are several causes of EBI, including mechanical injury^31^, vascular disruption^32^, and hippocampal injury^33^, among others. To explore the relationship between EBI and S100A9, we investigated whether the expression of S100A9 in the CSF of patients was increased by collecting CSF samples from patients within 24 h of onset (either before or after treatment). Using ELISA, we found that the concentration of S100A9 in the CSF of patients with SAH, although lower, was still generally higher than that in normal controls. Over time, it increased and finally leveled off. The S100A9 concentration is also correlated with the short-term prognosis of patients with SAH. These results indicate that there is a strong correlation between S100A9 and SAH. Previous studies have shown that microglial cells exhibit morphological changes in the first 24 h after intracranial vascular rupture^34^. Immunofluorescence staining was used to determine S100A9 distribution in different nerve cells in the brain tissue after SAH. The results showed that S100A9 was expressed more in microglia and less in other cells such as astrocytes and neurons, suggesting that microglia play an important role in connecting SAH with S100A9 expression. We observed that S100A9 knockout mice had improved neurological scores after SAH compared with WT mice. All learning, memory, and emotional functions of human beings are related to the hippocampus in the brain, and the hippocampus has strong plasticity^35^. H&E staining showed that S100A9 knockout mice had less brain edema and neuronal degeneration in the ipsilateral cortex and hippocampus. Apoptosis, programmed cell death, is a process that occurs in multicellular organisms and is characterized by cell shrinkage, nuclear fragmentation, and chromatin condensation^36^. These changes were observed using Nissl staining, especially in the ipsilateral cortex and hippocampus (CA1, CA3, and DG) after SAH. In addition, S100A9 knockdown significantly improved this phenomenon, as confirmed by TUNEL staining. After SAH, heme produced by the degradation of blood entering the subarachnoid space can activate microglia via TLR4 receptors, thereby inducing a proinflammatory cascade^37–39^. Therefore, we used hemin to construct an *in vitro* SAH model. Through the co-culture model of BV2 and HT22 cells, it was verified that the number of apoptotic cells reduced significantly after the addition of the S100A9 specific inhibitor, Paquinimod, indicating that Paquinimod has a protective effect on HT22 cells.

Neuroinflammation, mainly mediated by microglial activation and the recruitment of immune cells, is one of the key drivers of EBI after SAH^40^. Short-term prognosis after SAH can be improved by inhibiting neuroinflammation^41, 42^. Activation of microglia and M1 polarization after SAH were demonstrated by increased levels of iNOS, TNF-α, and IL-1β^43^. We further explored the relationship between S100A9 and inflammation by intracerebroventricular injection of the S100A9 recombinant protein. Our study showed that S100A9 knockdown reduced the expression of inflammatory factors in brain tissue after SAH, and the levels of inflammatory factors and microglia marker iba1 were increased again after the addition of S100A9 recombinant protein, suggesting that S100A9 aggravates neuroinflammation after SAH under certain conditions. NF-kB is an important nuclear transcription factor that responds to cells stress and cytokines. When the NF signaling pathway is activated by an upstream signal, the NF-kB complex p65 subunit is released and is further phosphorylated in the nucleus, combines with DNA, and it initiates transcription of inflammatory factors^44^. Our study showed that the levels of TLR4, MYD88, p-IκBα, p-p65, and nuclear translocation of p65 were increased after SAH, which could be inhibited by S100A9 knockdown or Paquinimod administration, and intervention with S100A9 recombinant protein could increase the expression levels of these genes again.

Our results confirmed that S100A9 knockout could ameliorate early brain injury after SAH by reducing neuroinflammation by inhibiting the TLR4/MYD88/NF-κB pathway activation. However, our study had certain limitations. In previous studies, S100A9 was shown to have different effects–proinflammatory or anti-inflammatory^45^, and we did not further explore the conditions under which S100A9 may manifest as an anti-inflammatory molecule after SAH. Second, the *in vivo* SAH process is complex, and the *in vitro* model we established cannot fully simulate it. Finally, whether S100A9 mediates inflammation via other receptor proteins has not been discussed. We only targeted the TLR4 receptor protein, hence, other pathway mechanisms remain to be elucidated.

## 5. Conclusion

In our study, S100A9 knockdown reduced early brain injury after SAH and improved short-term prognosis by inhibiting the TLR4/MYD88/NF-κB pathway activation, promoting microglial activation to M1, and reducing neuroinflammation. Therefore, S100A9 inhibitors may be therapeutic targets for alleviating early brain injury after SAH.

## Declarations

### Ethics approval and consent to participate

All animal experiments were approved by the Animal Experiment Center of Wuhan University.

### Consent for publication

Not applicable.

### Availability of data and materials

All raw data is provided as required.

### Authors’ contributions

GW, KH and ZZ jointly completed the experiment and wrote the original draft of this study. QT, YG and CL processed the data, ZL collected the data, GW, KH, ML and ZY designed the study, ML contributed to the project administration.

## Data Availability

All raw data is provided as required.

## Non-standard Abbreviations and Acronyms

SAH: subarachnoid hemorrhage
ELISA: enzyme-linked immunosorbent assay
TLR4: toll-like receptor 4
EBI: early brain injury
BBB: blood-brain barrier
PAMP: pathogen-associated molecular pattern
DAMP: damage-associated molecular pattern
PRRs: Pattern recognition receptors
HSP: heat-shock protein
HMGB1: high mobility group protein B1
RAGE: receptor for advanced glycation end products
PBS: phosphate buffer solution
MYD88: myeloid differentiation factor 88
IL-1β: interleukin-1β
TNF-α: tumor necrosis factor
WT: wild-type
Sham: sham-operated
PFA: paraformaldehyde
DAPI: 4-6-diamidino-2-phenylindole
ECL: enhanced chemiluminescence
CA: cornu smmonis
CSF: cerebrospinal fluid

## Acknowledgements

Thanks to Prof. Mingchang Li for funding this study.

## Author details

^1^Department of Neurosurgery, Renmin Hospital of Wuhan University, Wuhan 430060, Hubei Province, China. ^2^Department of Critical Care Medicine, Renmin Hospital of Wuhan University, Wuhan 430060, Hubei Province, China. ^3^Department of Rehabilitation Medicine, Renmin Hospital of Wuhan University, Wuhan 430060, Hubei Province, China.

